# Citywide Nucleic Acid Screening of SARS-CoV-2 Infections in Post-lockdown Wuhan, China: Results and Implications

**DOI:** 10.1101/2020.06.29.20142554

**Authors:** Shiyi Cao, Yong Gan, Chao Wang, Max Bachmann, Yuchai Huang, Tiantian Wang, Liqing Li, Kai Lu, Heng Jiang, Yanhong Gong, Hongbin Xv, Xin Shen, Qingfeng Tian, Chuanzhu Lv, Fujian Song, Xiaoxv Yin, Zuxun Lu

## Abstract

**Background:** After the outbreak of Coronavirus disease in 2019 (COVID-19), stringent lockdown measures were imposed in Wuhan between January 23, 2020 and April 8, 2020. To provide evidence on the post-lockdown risk of COVID-19 epidemic in Wuhan, the city government conducted a citywide nucleic acid screening of SARS-CoV-2 infection between May 14 and June 1, 2020.

**Methods:** All city residents aged ≥6 years were potentially eligible to participate the screening programme. The rate of detection of asymptomatic infected cases was calculated, and their demographic and geographic distributions were investigated. ArcGIS 10.0 was used to draw a geographic distribution of asymptomatic infected persons.

**Results:** The screening programme recruited a total of 9,899,828 persons (response rate, 92.9%). The screening found no newly confirmed patients with COVID-19, and identified 300 asymptomatic infected cases (detection rate 0.303/10,000). In addition, 107 of 34,424 previously recovered patients with a history of COVID-19 diagnosis were tested positive (relapse rate, 0.31%). Virus culture of SARS-CoV-2 was negative for all 300 asymptomatic cases and all 107 recovered COVID-19 patients. A total of 1,174 close contacts of asymptomatic cases were traced and all of them had a negative nucleic acid testing result.

**Conclusions:** Prevalence of COVID-19 nucleic acid test positivity was very low in the Wuhan general population, in recovered cases and in contacts of asymptomatic cases, five to eight weeks after the end of lockdown. These findings help resolve concerns about the post-lockdown risk of COVID-19 epidemic, and promote the recovery of economy and normal social life in Wuhan.

## Introduction

The Coronavirus Disease 2019 (COVID-19) was first reported in December 2019, and has spread to more than 200 countries and regions. Following strict lockdown measures, the COVID-19 epidemic was generally under control in China, and progressed into a post-lockdown phase, similar to South Korea, Italy, Switzerland, and other countries. However, China faces new problems and challenges, including how to accurately assess the current risk of the COVID-19 epidemic, how to avoid new waves of outbreaks of COVID-19 epidemic, and how to facilitate the resumption of economy and normal social life. Wuhan was the city most severely affected by COVID-19 in China. Wuhan had been locked down on January 23, 2020 and unblocked on April 8 after the effective control of COVID-19 epidemic. Over the past two months, only a few sporadic COVID-19 cases have been found. However, people still were very worried about the COVID-19 epidemic in Wuhan, which seriously affected the resumption of industrial production and the wider economy, and hampered the normal life of residents. In order to ascertain the current status of the COVID-19 epidemic, the government of Wuhan carried out a comprehensive citywide nucleic acid screening of SARS-CoV-2 infection from May 14, 2020 to June 1, 2020.

This study reports the process and results of this citywide nucleic acid screening, and discusses its significance, providing references for other countries and regions in the post-lockdown phase to accurately prevent and control the COVID-19 epidemic, resolve concerns, and promote the recovery of society and economy.

### Testing process and methods

### Targeted population

Wuhan has about 11 million residents in total, with 7 urban and 8 suburban districts. Residents are living in 7,280 residential communities (or residential enclosures, “xiao-qu”), and each residential community could be physically isolated from other communities for the purpose of preventing transmission of COVID-19.

The screening programme included participants who were aged ≥ 6 years, and provided a written informed consent after reading a statement that explained the purpose of the testing. All residents currently living in Wuhan were eligible, including those with a history of COVID-19 diagnosis.

### Organizational guarantee and community mobilization

A citywide nucleic acid testing group was established, with specialized task teams contributing to comprehensive coordination, technical guidance, quality control, information management, communication, and supervision of testing. The government invested 900 million yuan (RMB) in the testing programme, which involved 63 nucleic acid testing institutions, 1,451 healthcare workers and 701 testing equipment. Public information communication was implemented through mass media, mobile messages, WeChat groups, and residential community broadcasts, so as to increase residents’ awareness and the participation. Door-to-door testing was provided for residents if they had difficulty with mobility. All detected asymptomatic infected persons were isolated for two weeks in designated hotels managed by health professionals from community medical centres, and their close contacts were traced.

### Sampling, testing and quality control

Trained personnel collected nasopharyngeal and pharyngeal samples. All sampling personnel received the standard training. To minimise the risk of cross-infection, the sampling process strictly followed a disinfection process and ensured environmental ventilation. Real-time reverse transcriptase-polymerase chain reaction (RT-PCR) assay method was used for the nucleic acid testing. Samples were tested within 24 hours of collection, and any samples that could not be tested timely were stored at -70°C or below. Two trained staff recorded sample information into a specifically designed database. In addition to “single testing” (separately testing of a single individual sample), “mixed testing” was also performed for some samples to increase the efficiency, in which no more than 5 samples were mixed in equal amounts, and tested in the same test tube. If a mixed sample was tested positive for COVID-19, all mixed samples were recollected again and they were separately tested.^1^ The colloidal gold antibody test was also performed for asymptomatic infected persons.

### Data collection

Before sample collection, residents electronically (using a specifically designed smartphone app) self-uploaded their personal information, including ID number, name, sex, age, and place of residence. Then trained staff interviewed them on the history of the COVID-19 infection and previous nucleic acid testing. All information was entered into a central database. The testing results were continuously uploaded to the central database by testing laboratories. The pre-existing unique identification code for each resident, was used as the programme’s identification number, to ensure information accuracy during the whole process of screening, from sampling, nucleic acid testing, result reporting, isolation of asymptomatic infected cases and tracing of their close contacts. Contact tracing investigations were conducted on participants who tested positive for SARS-CoV-2, to track and manage their close contacts. All screening information was kept strictly confidential and was not allowed to be disclosed or used for other purposes rather than clinical and public health management. Personal information of asymptomatic infected persons was only disclosed to designated medical institutions and community health centres for the purpose of medical isolation and identification of close contacts.

### Biological security guarantee

Nucleic acid testing was performed in second-level biosafety laboratories. Sampling and testing personnel adopt the personal protective measures according to the standard of biological third-level safety protection laboratories. The laboratory put control measures in place to guarantee biological safety in accordance with relevant regulations.^2^

### Result query and feedback

Two to three days after the nucleic acid screening, participants could inquire the test results using the WeChat or Alipay app through their unique ID numbers. The results included coloured health codes and text descriptions of the nucleic acid testing. When the test result was negative, the health code was green; otherwise, the health code was red.

### Definition of confirmed cases, asymptomatic infected person and close contacts

In this study, confirmed COVID-19 cases were previously diagnosed by designated medical institutions. Asymptomatic infected person referred to those who had no history of COVID-19 diagnosis or a positive nucleic acid test, but they tested positive for SARS-CoV-2 and did not show any clinical symptoms at the time of nucleic acid testing. Close contacts were those who socially interacted with suspected or infected cases within 2 days before symptom onset, or closely contacted with an asymptomatic infected person within 2 days before the nucleic acid sampling.^3^ Relapse rate in this study was defined as the proportion of participants with a positive nucleic acid testing result in previously recovered COVOD-19 patients.

### Data analysis

Because the estimated detection rates were extremely low, we calculated 95% confidence intervals of estimated proportions using Pearson-Klopper exact method, implemented through R package “binom” version 1.1-1. SPSS version 22.0 was used for other statistical analyses. We analyzed the distribution of participants and asymptomatic infected persons and assessed the spearman correlation between the detection rate of asymptomatic infected persons and the prevalence of confirmed COVID-19 cases. Differences in detection rate of asymptomatic cases by sex and age group were assessed using the *χ*^2^ test. ArcGIS 10.0 was used to draw a geographic distribution of asymptomatic infected persons. A value of *P*<0.05 (two-tailed) was considered statistically significant.

## Results

There were 10,652,513 eligible people aged ≥6 years in Wuhan (94.1% of the total population). The nucleic acid screening was completed in 19 days (from May 14, 2020 to Jun 1, 2020), and recruited a total of 9,899,828 persons (participation rate, 92.9%). The majority of non-participants were not in Wuhan at the time of the screening.

The screening found no confirmed COVID-19 cases, and identified 300 asymptomatic infected cases with a detection rate of 0.303/10,000 (95% CI 0.270 – 0.339). A total of 1,174 close contacts of the asymptomatic cases were traced, and they all tested negative for the COVID-19. There were 34,424 previously diagnosed and recovered COVID-19 cases (accounting for 68.4% of recovered patients) participated in the screening. Of the 34,424 participants with a history of COVID-19, 107 were tested positive, and giving a relapse rate of 0.310% (95% CI 0.423 −0.574%).

Virus culturing were negative for all samples of asymptomatic cases and those with a history of COVID-19 diagnosis, indicating no “viable virus” in those with a positive result of nucleic acid testing in this study.

In this screening programme, 76.7% samples were single testing and 23.3% were mixed testing. The detection rate in asymptomatic cases was 0.334/10,000 and 0.242/10,000, respectively, through the two testing approaches.

Among the 300 asymptomatic infected persons, there were 132 males (0.256/10,000) and 168 females (0.355/10,000). They aged from 10 to 89 years. The lowest detection rate was 0.124/ 10,000 (12 cases) in children or adolescents aged 17 and below, and the highest detection rate was 0.442/10,000 (88 cases) among the elderly aged 60 years and above (see Table 1).

**Table 1.**
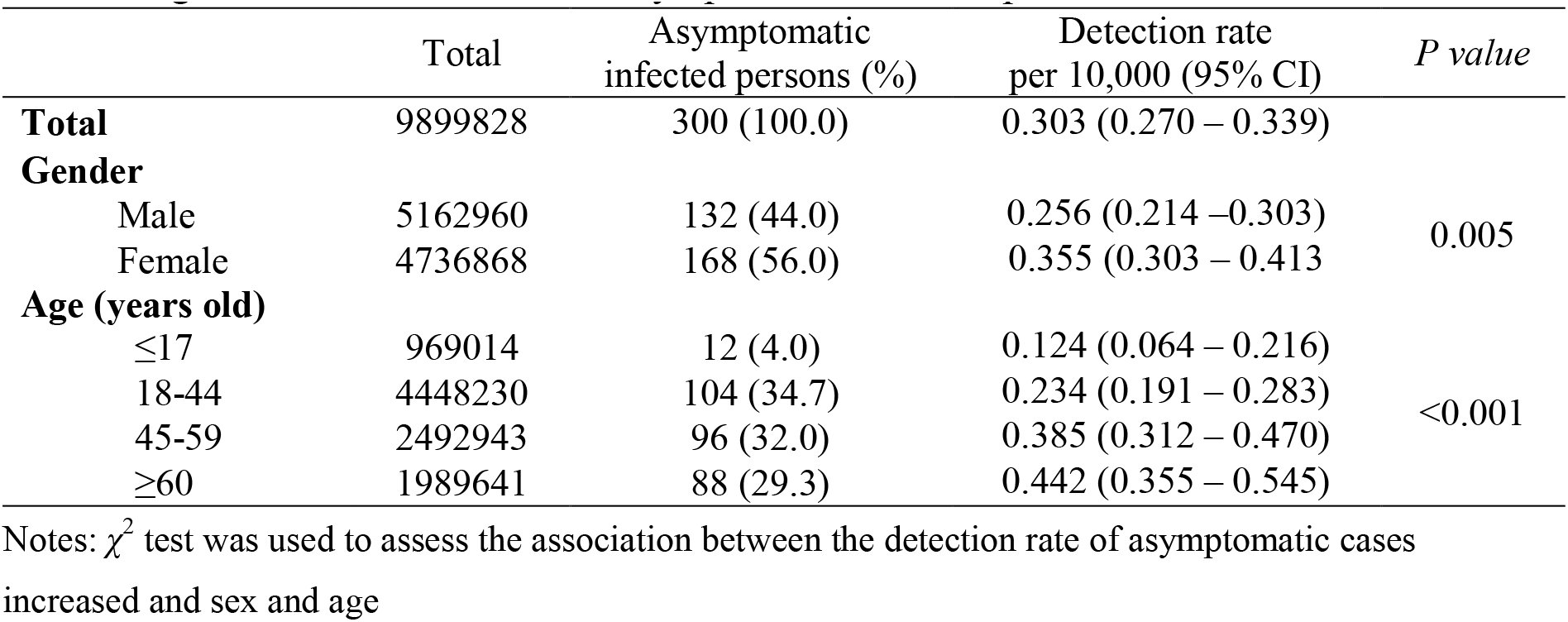
Age and sex distributions of asymptomatic infected persons.

|  | Total | Asymptomatic<br>infected persons (%) | Detection rate<br>per 10,000 (95% CI) | *P value* |
| --- | --- | --- | --- | --- |
| **Total** | 9899828 | 300 (100.0) | 0.303 (0.270 – 0.339) |  |
| **Gender** |  |  |  |  |
| Male | 5162960 | 132 (44.0) | 0.256 (0.214 – 0.303) | 0.005 |
| Female | 4736868 | 168 (56.0) | 0.355 (0.303 – 0.413) |  |
| **Age (years old)** |  |  |  |  |
| ≤17 | 969014 | 12 (4.0) | 0.124 (0.064 – 0.216) | <0.001 |
| 18-44 | 4448230 | 104 (34.7) | 0.234 (0.191 – 0.283) |  |
| 45-59 | 2492943 | 96 (32.0) | 0.385 (0.312 – 0.470) |  |
| ≥60 | 1989641 | 88 (29.3) | 0.442 (0.355 – 0.545) |  |

Notes: $\chi^2$ test was used to assess the association between the detection rate of asymptomatic cases increased and sex and age

The three occupations with the most detected cases were domestic and unemployed (24.3%), retired (21.3%), and public service workers (11.7%) (see Figure 1).

**Figure 1.**
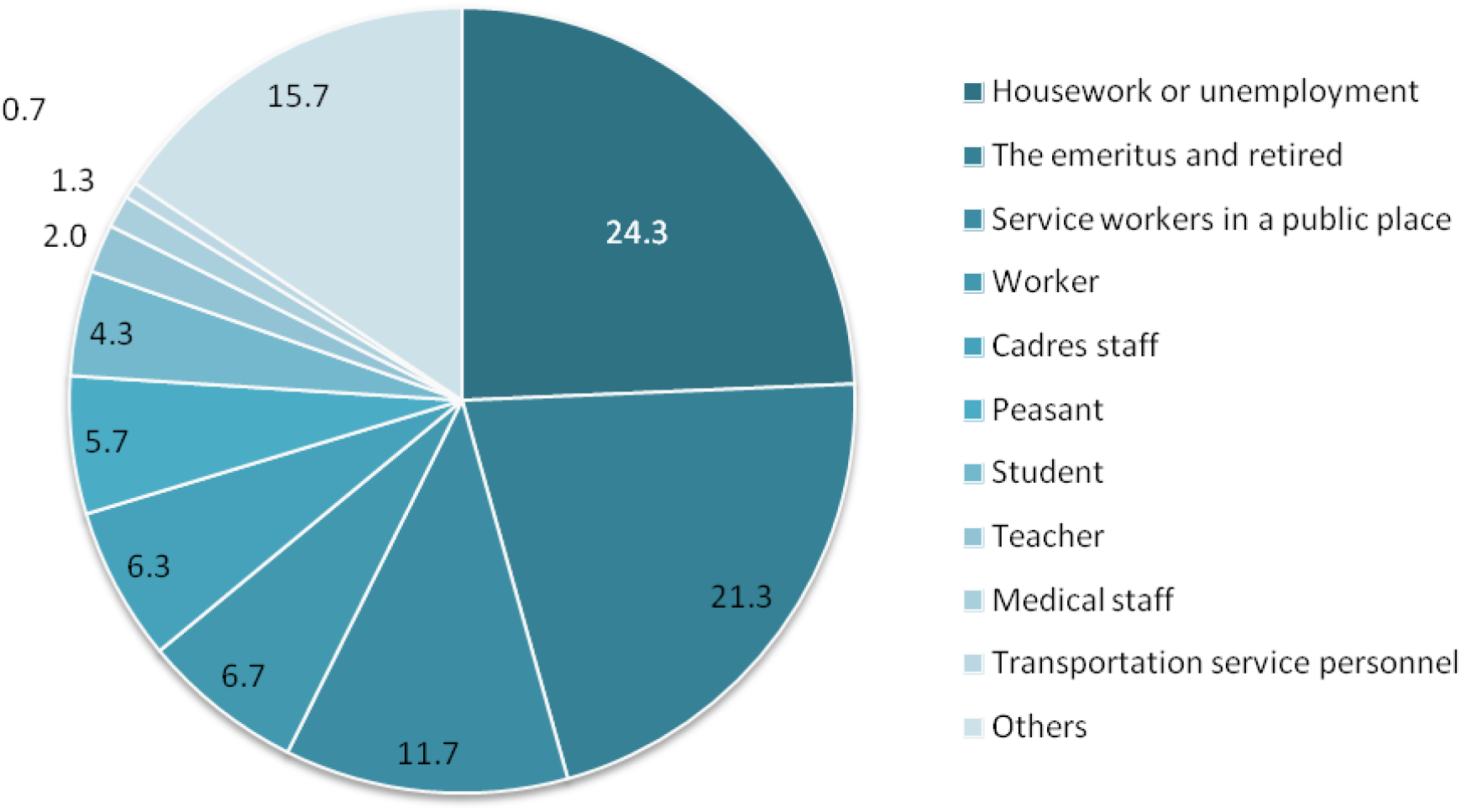
The occupation distribution of asymptomatic infected patients (%) Notes: Others included the self-employed, soldier and so on)

The detection rate of asymptomatic infected persons in urban districts was 0.442/10,000. Specifically, the detection rate was 0.741/10,000 in Wuchang district, 0.555/10,000 in Qingshan district, and 0.531/10,000 in Qiaokou district. A lower detection rate was found in suburban districts (Caidian district, Jiangxia district, Huangpi district, Dongxihu district, and Xinzhou district) (47 cases, 0.132/10,000) (Figure 2).

**Figure 2.**
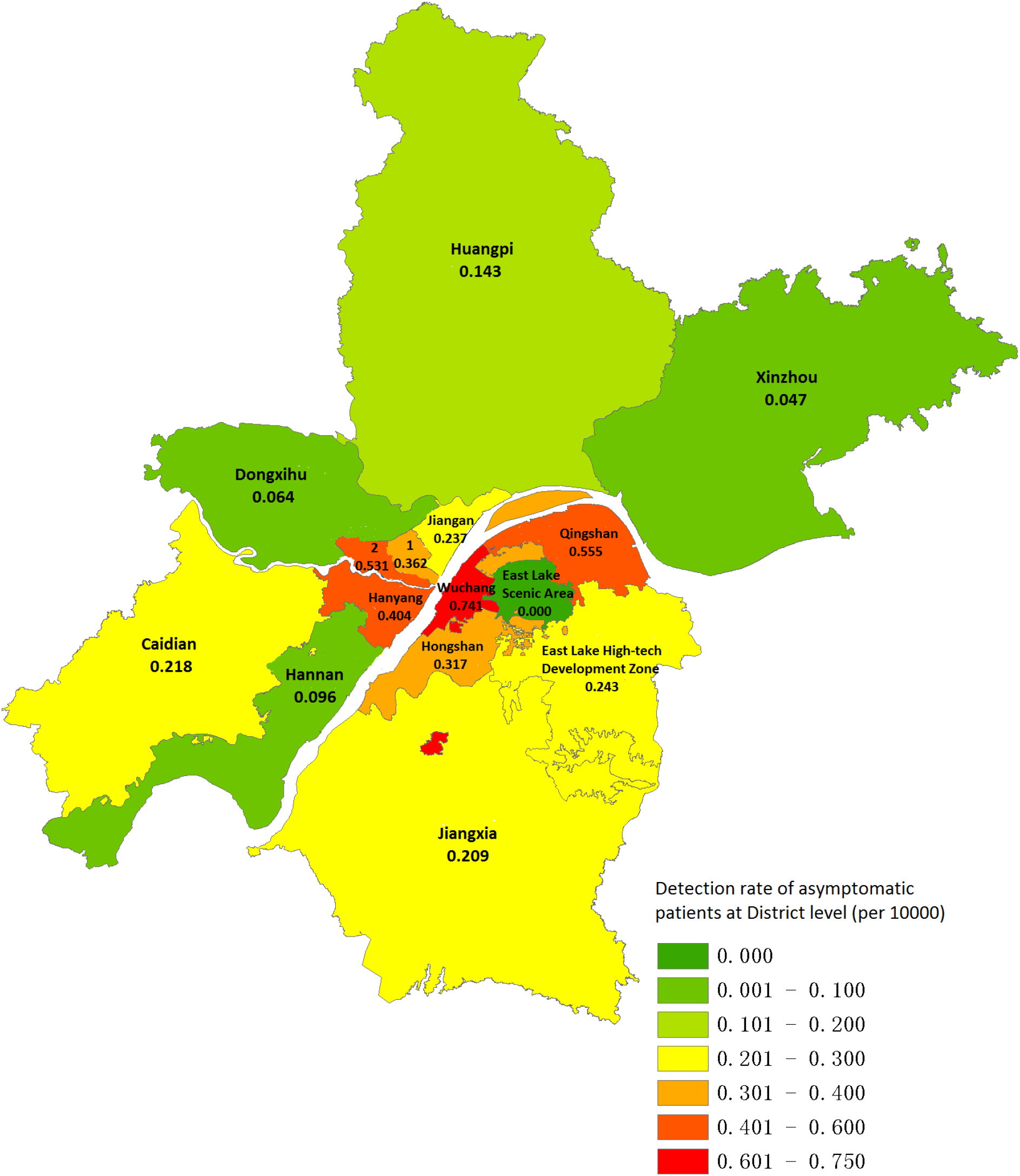
The geographic distribution of the detection rate of asymptomatic infected patients (Notes: 1 represents Jianghan district; 2 represents Qiaokou district)

Among 7,280 residential communities, asymptomatic infected persons were identified in 265 (3.6%) communities (only one case was detected in 246 communities), while no asymptomatic infected cases were found in the rest communities.

Among asymptomatic infected persons, 63.3% (n=190) were IgG (+), indicating that most asymptomatic infected persons were previously infected. The proportion with both IgM (-) and IgG (-) was 36.7% (n=110), indicating the possibility of infection window or false positive results of the nucleic acid testing.

A higher detection rates of asymptomatic infected persons were in Wuchang, Qingshan and Qiaokou districts, and the prevalence of previously confirmed COVID-19 cases were 68.243/10,000, 53.767/10,000, and 100.047/10,000, respectively, in the 3 districts. Districts with a high detection rate of asymptomatic infected persons generally had a higher prevalence of previously diagnosed infections (*r* =0.710, *P*=0.003) (Figure 3).

**Figure 3.**
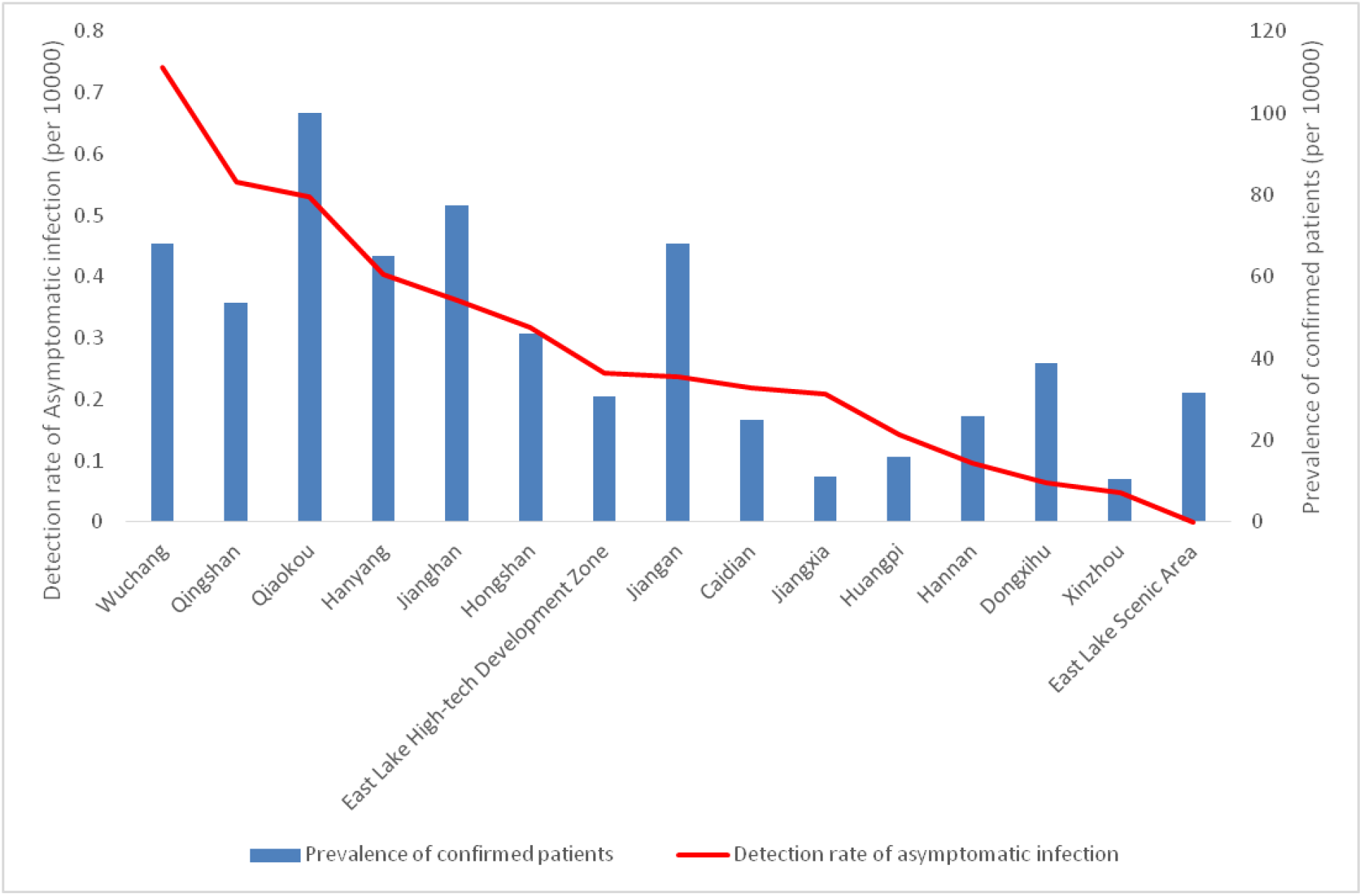
The correlation between the detection rate of asymptomatic infected patients and the prevalence of previously confirmed patients with COVID-19 in each district in Wuhan

## Discussion

### The prevention and control of COVID-19 epidemic is effective in Wuhan

The citywide nucleic acid screening of SARS-CoV-2 infection in Wuhan recruited nearly 10 million people, and found no new confirmed (that is, previously diagnosed) cases with COVID-19. The detection rate of asymptomatic infected persons was very low, and there was no evidence of transmission from asymptomatic infected persons to traced contacts. No asymptomatic infected cases were identified in 96.4% of residential communities, indicating that Wuhan has made remarkable achievements in control of the COVID-19 epidemic.

### Infectivity of identified asymptomatic infected cases

Previous studies have shown that the asymptomatic infected patients with COVID-19 have some infectivity.^4^ Some asymptomatic infected patients might become symptomatic confirmed cases.^5^ Compared with the symptomatic patients, asymptomatic infected persons generally have low quantity of viral loads and a short duration of viral shedding, which decrease the transmission risk of SARS-CoV-2.^6^ Virus culturing was carried out on samples from asymptomatic infected persons, and no viable SARS-CoV-2 virus was found. All close contacts tested negative, indicating that the asymptomatic infected persons were not infectious. Even so, asymptomatic infected persons have been under strict centralized isolation and medical observation, in order to block any possible risk of virus transmission.

### There is a low relapsing rate in recovered patients with COVID-19

There was a low relapsing rate in recovered COVID-19 patients in Wuhan. However the current study found that recovered patients with positive results of nucleic acid testing were not infectious. Besides the nasopharyngeal and pharyngeal swabs, nucleic acid can be detected in sputum, stool, urine and tears in the samples of confirmed cases,^7HYPERLINK \l “bookmark7”, 8^ although the clinical and public health implications of such positive results remain unclear. In order to minimise the possible risk of relapsing, it is necessary to follow up recovered COVID-19 cases regularly.^9^

### The geographic distribution of asymptomatic cases is consistent with the distribution of previously confirmed cases

Considering the strong force of infection of COVID-19 ^10-12^, it is expected that the number of confirmed cases is associated with the risk of being infected in communities. With the centralized isolation and treatment of all COVID-19 cases during the lockdown period in Wuhan, the risk of residents being infected in the community has been greatly reduced. When susceptible residents are exposed to a low dose of virus in the air and on surfaces, they may tend to be asymptomatic due to their own immunity, instead of showing clinical symptoms of the COVID-19.^10^ We found that the detection rates of asymptomatic infected persons in different districts of Wuhan were correlated with the prevalence of previously confirmed cases. This was in line with the temporal and spatial evolution (especially the long-tailed characteristic) of infectious diseases, and, to some extent, supports the accuracy of the previous epidemic report in Wuhan.

### This study provides population-based clues to explain the decreases of viral load and transmission shown previously

Previous laboratory virus culture and genetic studies^11, 12^ showed that the virulence of the COVID-19 may be gradually weakening, and the newly infected persons were more likely to be asymptomatic and with a lower viral load than in earlier reports. The citywide nucleic acid screening programme in Wuhan provides additional evidence regarding the virulence and transmission of SARS-CoV-2 after relaxing lockdown measures. Nonetheless, it is too early to be complacent, because most people in Wuhan are still susceptible to COVID-19 and preventive measures, including safe social distancing, should be sustained. Especially, vulnerable populations with weakened immunity or co-morbidities, or both, should continue to be appropriately shielded.

### Global reference value

The citywide nucleic acid screening result shows that the COVID-19 epidemic has been brought under control in Wuhan. This helps to alleviate residents’ anxiety about COVID-19 in Wuhan and to facilitate the recovery of the economy and normal social life. The successful implementation of this citywide nucleic acid screening in Wuhan provides helpful experience to other countries and regions entering the post-lockdown phase. However, some limitations of this study need be pointed out. First, this was a cross-sectional screening programme, and we are unable to assess the changes over time, including changes in the prevalence of asymptomatic infected persons, mental health of residents, and social and economic development. Second, large scale citywide screening required a certain financial support and it may not be feasible in less-developed countries and regions. Third, the implementation of the citywide screening requires the involvement of all residents. Residents need to have good health literacy, awareness of prevention and control measures, and willingness of participation. Finally, although a positive result of nucleic acid testing certainly reveals the existence of the viral DNA, the possibility of false negative results could not be completely ruled out, in particular due to low level of virus loads or inadequate collection of samples.^13^ However, even if test sensitivity was as low as 50%, then the actual prevalence would be twice as high as reported in this study, but would still be very low.

In summary, the citywide nucleic acid screening of SARS-CoV-2infection in post-lockdown Wuhan shows the determination and perseverance of the government and residents. The detection rate of asymptomatic cases was very low (0.303/10,000), and there was no evidence that the identified asymptomatic cases were infectious. It helps resolve concerns about the post-lockdown risk of the COVID-19 epidemic and facilitate the recovery of economy, and reopening of educational and medical institutions in Wuhan. It also provides evidence to enable the government to adjust the prevention and control strategies to recover the outpatient and inpatient departments of medical institutions, to meet the healthcare needs of residents with other health conditions and mental health. Further studies are required to fully evaluate the impacts and cost-effectiveness of the citywide screening of SARS-CoV-2 infections on population’s health, health behaviours, economy, and society.

## Data Availability

All data referred to in the manuscript can be available if required.

## Acknowledgements

We would like to thank all institutions and all citizens in Wuhan for their support for testing work. We also would like to thank the big data and investigation group of COVID-19 prevention and control institution in Wuhan (the data and investigation group of Wuhan Municipal Health Commission) for their efforts in the data collection.

## Authors’ Contributions

SYC, CW, XXY, and ZXL conceived the study. CW, YCH, TTW, KL, HBX, and XS participated in the acquisition of data. YCH, TTW, and LQL analyzed the data. HJ, YHG, and FJS gave advice on methodology. QFT and CZL investigated the responses to the citywide nucleic acid testing among residents lived in outside of Wuhan city. SYC, YG, CW, XXY drafted the manuscript, YG, MB, and FJS revised the manuscript, and MB and FJS critically commented and edited the manuscript. All authors read and approved the final manuscript. ZXL is the guarantor of this work and, as such, had full access to all the data in the study and takes responsibility for the integrity of the data and the accuracy of the data analysis.

